# Using natural language processing to study homelessness longitudinally with electronic health record data subject to irregular observations

**DOI:** 10.1101/2023.03.17.23287414

**Authors:** Alec B. Chapman, Daniel O. Scharfstein, Ann Elizabeth Montgomery, Thomas Byrne, Ying Suo, Atim Effiong, Tania Velasquez, Warren Pettey, Richard E. Nelson

## Abstract

The Electronic Health Record (EHR) contains information about social determinants of health (SDoH) such as homelessness. Much of this information is contained in clinical notes and can be extracted using natural language processing (NLP). This data can provide valuable information for researchers and policymakers studying long-term housing outcomes for individuals with a history of homelessness. However, studying homelessness longitudinally in the EHR is challenging due to irregular observation times. In this work, we applied an NLP system to extract housing status for a cohort of patients in the US Department of Veterans Affairs (VA) over a three-year period. We then applied inverse intensity weighting to adjust for the irregularity of observations, which was used generalized estimating equations to estimate the probability of unstable housing each day after entering a VA housing assistance program. Our methods generate unique insights into the long-term outcomes of individuals with a history of homelessness and demonstrate the potential for using EHR data for research and policymaking.

## Introduction

Access to stable housing is an important social determinant of health (SDoH) and is linked to several important health outcomes and general well-being. In recent years, Electronic Health Records (EHR) have increasingly been utilized as a data source for studying homelessness and other adverse SDoH.^1–10^ Many of these studies have utilized natural language processing (NLP), a set of techniques for extracting information from unstructured clinical texts. Improved measurement of housing instability offers a number of benefits to researchers and policymakers such as the ability to study risk factors for becoming homeless or the effectiveness of homelessness interventions.

One advantage of using clinical data to study housing instability is the longitudinal nature of the EHR. Housing instability is not only a point-in-time event, and sustained access to stable housing is an important clinical outcome for patients with a history of homelessness. Studying longitudinal housing requires repeated measures of housing status and long-term follow up, presenting a challenge to researchers examining long-term housing outcomes and an opportunity for leveraging the temporal structure of EHR data.

A challenge in using EHR data for studying longitudinal housing outcomes is irregular observation. Ideal longitudinal studies measure outcomes at regular, fixed intervals for each subject for the entire study period. Clinical notes and other EHR documentation, on the other hand, are recorded only when a patient has a healthcare encounter and a clinician documents information about their housing status. This results in highly irregular encounters and a large amount of incomplete information. Furthermore, patients who are unstably housed may have more information recorded in the EHR, as they may be more likely to utilize services related to housing instability and clinicians may be more likely to record instances of housing instability than cases where a patient is stably housed. It could also be the case that patients who are unstably housed may have less information recorded in the EHR, due to their current living conditions preventing them from seeking care for their health needs. Either scenario can lead to “informative assessment times,” which occur when there is an imbalance in the frequency of observations for stably housed and unstably housed individuals and can lead to biased estimates of housing instability.^11–14^

This study aimed to address these challenges by developing a framework for studying housing instability longitudinally using EHR data. We applied these methods in the setting of a longitudinal follow up of patients enrolled in a housing assistance program in the Department of Veterans Affairs (VA). First, we used a previously validated NLP system to extract housing status over a three-year period following program entry. Second, we fit a recurrent event model to calculate inverse intensity weights used to adjust for the irregularity of patient visits. Third, we fit marginal regression models to estimate the probability of being unstably housed each day over the three-year period.

## Methods

### Data and Setting

We performed our analysis using data from a cohort of 35,074 Veterans who were enrolled in Supportive Services for Veteran Families (SSVF) and received care in VA. SSVF is a program that provides support to Veterans who are homeless or at risk of becoming homeless. Veterans enrolled in SSVF may receive several forms of assistance including case management, housing search assistance, and direct payments for housing-related costs in the form of temporary financial assistance (TFA). Previous studies have evaluated the association between patient characteristics and different forms of treatment with housing status when exiting SSVF.^15,16^ However, studies examining the long-term housing outcomes for SSVF participants have been limited.

Treatment information and other baseline characteristics were retrieved from the Homeless Management Information System (HMIS),^*^ a national database for programs receiving federal assistance from HUD. HMIS variables included what forms of assistance the patient received, the patient’s entry and exit date into SSVF, patient demographics, and other baseline characteristics. We next linked HMIS data to patient records in VA Corporate Data Warehouse (CDW), a clinical data repository containing data collected from Veteran encounters including patient demographics, administrative data, and clinical notes.

### Extracting housing status from the EHR

Housing status was extracted from clinical notes in the EHR using a previously validated NLP system called ReHouSED ^4^. Briefly, ReHouSED is a rule-based NLP system that takes as input a clinical note and extracts phrases representing concepts related to housing status, assigns linguistic attributes such as negation and temporality, and infers a document-level classification that represents the overall housing status of the Veteran according to the note. Notes are assigned one of three document classifications: “Stable,” indicating explicit evidence of stable housing; “Unstable,” indicating explicit evidence of unstable housing; or “Unknown,” meaning that the note lacked explicit documentation of housing status and a classification could not be determined. ReHouSED was implemented in Python 3.7.4 using medspaCy 0.0.1.4.^17^ We processed all notes containing keywords related to housing from 180 days prior to a Veteran’s entry into SSVF up to three years (1,095 days) after. For each Veteran, we aggregated to a housing-related encounter level (i.e., a day that a Veteran presented to VA for care and had information on housing documented in a note) by classifying a Veteran as unstably housed if half or more of the notes on a day were classified as “Unstable” after excluding “Unknown” notes.

### Independent variables

We extracted baseline covariates from HMIS and CDW as in previous studies of SSVF.^15,16^ For each patient, we identified patient demographics such as age, race, gender, and marital status; socioeconomic indicators such as employment, education, and income; SSVF treatment information such as whether they received case management or temporary financial assistance (TFA); whether the patient received other social services such as Supplemental Nutrition and Assistance Program (SNAP) or disability benefits; insurance type (e.g., Medicare, Medicaid, or private insurance); whether the patient received services from other VA homelessness programs such as Housing and Urban Development-VA Supportive Housing (HUD-VASH); clinical comorbidities including mental health diagnoses and the patient’s Charlson Comorbidity Index;^18^ inpatient and outpatient cost in the previous 365 days; and the patient’s housing history, including whether the patient was homeless or merely at risk of becoming homeless at the time of SSVF entry and the number of times homeless in the previous 3 years. Additionally, we included NLP-extracted measures of long-term and short-term housing history, namely the number of housing-related encounters in the 180 days before entering SSVF and the 14 days before. After adding an indicator for variables with high amounts of missingness, we restricted the analysis to observations with no missing covariates.

#### Statistical Analyses and Models

All statistical analyses were performed using R 4.1.2. Code and supplemental materials are available on GitHub.^†^

### Descriptive statistics

We first plotted the observed probability of unstable housing before and after entering SSVF by calculating the proportion of patients each day who were classified as “Unstable” out of all patients assigned a classification. Next, we plotted the counts of housing-related encounters each day. We also calculated summary statistics describing the frequency and spacing of encounters across individuals. To assess the degree of follow-up, we identified the final observation for each patient and plotted the proportion of patients who had at least one visit at or after each time point.

### Inverse intensity weighting to adjust for encounter frequency

Next, we implemented inverse intensity weighting (IIW) to adjust for irregularity in the observation of housing outcomes. As housing status was extracted from notes recorded in the EHR, the outcome variable was only observed when a patient presented for care and housing status was documented in a note, which occurred at highly irregular intervals. Furthermore, we hypothesized that patients who were unstably housed were more likely to visit the VA and have documentation of their housing status, which can lead to bias due to imbalance in the frequency of observation.^11,12,14^ IIW, proposed by Lin et al.,^13 21^adjusts for imbalance in visit frequency by fitting a recurrent event model and weighting visits by the inverse of the predicted risk of having an encounter on that day (i.e., the intensity). IIW assumes that patients are “assessed at random” (AAR), which states that a patient’s risk of an encounter at a given time *t* is independent of their current and future housing status conditional on their baseline covariates, housing history up until but not including time *t*, and other observed time-varying auxiliary variables. Patient encounters that occur with high intensity are weighted lower in outcome models, while encounters that occur with lower intensities are weighted higher, resulting in a pseudo-population in which an individual’s risk of an encounter at a given time is independent of their housing status. IIW is implemented in the R package *IrregLong* using a proportional intensity model.^‡^

The primary variables included in the intensity model were time-varying variables related to the encounter process, such as the patient’s last observed housing status (i.e., the NLP classification of their last housing-related encounter); whether they were admitted to an inpatient ward during their previous encounter; and whether they were enrolled in SSVF on the previous day. We also included patients’ baseline characteristics, SSVF treatment information, and NLP-derived measures of housing history. After fitting the proportional intensity model, a patient’s inverse intensity weight is calculated as the inverse of their predicted intensity at time 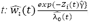, where Z_i_(t) is the vector of baseline covariates and time-varying auxiliary variables at time *t*, 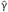 is a vector of regression coefficients, and 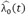 is a constant baseline hazard. Following Bůžková and Lumley,^19^ *IrregLong* uses stabilized weights that multiplies the inverse intensity weight by the baseline hazard, so that the final weights are: 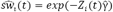.

### Outcome regression models

Next, we estimated the probability of unstable housing each day from 180 days before entry up to 1,095 days after using inverse-intensity weighted generalized estimating equations (GEE) with a logit link function. Encounters were clustered by patient using an independence working correlation structure. We represented time using natural cubic splines with knots at the following pre-defined follow up times: the day of entry, 30 days after entry, 60 days, 90 days, 180 days, 1 year, and 2 years. We chose these time points because we hypothesized that there would be a larger change in probability of unstable housing immediately following entry to the program and less variation in the long term. 95% confidence intervals were calculated 1000 bootstrap iterations. To evaluate the impact of IIW on estimates of housing instability, we fit a second unweighted GEE model.

## Results

### Descriptive statistics

Of the 35,074 patients included in our cohort, 211 were dropped due to missing covariates and 2,541 did not have any encounters in three-year follow up period and did not contribute to fitting the longitudinal models. **Table 1** displays summary statistics for demographics of the remaining 32,322 patients included in the final analysis.

**Table 1.** Patient demographics.

| Characteristic | N = 32,322 (%) |
| --- | --- |
| Age (mean, SD) | 50.7 (12.9) |
| <40 y | 7,700 (24%) |
| 40 to <50 y | 5,049 (16%) |
| 50 to <60y | 10,750 (33%) |
| >=60 y | 8,823 (27%) |
| Sex |  |
| Male | 27,950 (86%) |
| Female | 4,071 (13%) |
| Missing | 301 (0.9%) |
| Race |  |
| White | 17,289 (53%) |
| Black/African American | 14,164 (44%) |
| American Indian/Alaska Native | 456 (1.4%) |
| Other/Missing | 413 (1.3%) |
| Total monthly income |  |
| 0 | 9,378 (29%) |
| >0 to 500 | 3,208 (9.9%) |
| >500 to 1500 | 14,118 (44%) |
| >1500 | 5,618 (17%) |
| Education |  |
| Less than high school | 633 (2.0%) |
| High school diploma | 8,700 (27%) |
| Some college | 4,757 (15%) |
| College degree | 2,974 (9.2%) |
| Missing/Unknown | 15,258 (47%) |
| Family |  |
| Spouse or partner | 5,940 (17%) |
| Children | 6,673 (20.1%) |
| Insurance |  |
| Medicaid | 4,176 (13%) |
| Medicare | 2,573 (8.0%) |
| VA medical history |  |
| Mental health diagnosis | 20,159 (62%) |
| VA outpatient costs in last year (mean, SD) | 10,073 (12,634.3) |
| VA inpatient costs in last year (mean, SD) | 8,091.5 (28,675.7) |
| Homeless in last 3 years | 20,097 (62%) |
| SSVF services |  |
| Rapid rehousing (RR) | 20,943 (65%) |
| Homelessness prevention (HP) | 9,673 (30%) |
| RR + HP/Missing | 1,696 (5.2%) |
| Temporary financial assistance | 22,686 (70%) |
| Case management | 25,917 (80%) |
| Other VA homelessness programs |  |
| HUD-VASH | 6,841 (21%) |
| GPD | 4,061 (13%) |
| Other | 767 (2.4%) |

**Figure 1** shows the observed daily prevalence of unstable housing (i.e., the proportion of patients classified as unstable out of all patients with a housing-related encounter on that day) the counts of observed encounters, and the proportion of patients remaining in the analysis each day. Housing instability appeared to increase during the 180 days before entering SSVF, with a sharp decline beginning approximately 30 days before program entry. This trend continued for approximately 90 days after entering the program, after which there was a slight increase throughout the remainder of the study period. There were a total of 1,164,402 housing-related encounters, with encounters being most frequent in the time surrounding SSVF entry but with a large number of encounters throughout the entire study period. During those encounters, there were a total of 1,530,758 notes classified by the NLP as “stable” (49%) or “unstable” (51%). Over 75% of patients had an encounter in the final year of the study period, and 50% had an encounter in the last 989 days. Patients had a mean of 36 (median 20) encounters after entering SSVF, with an average of 21.7 (median 7) days between encounters.

**Figure 1.**
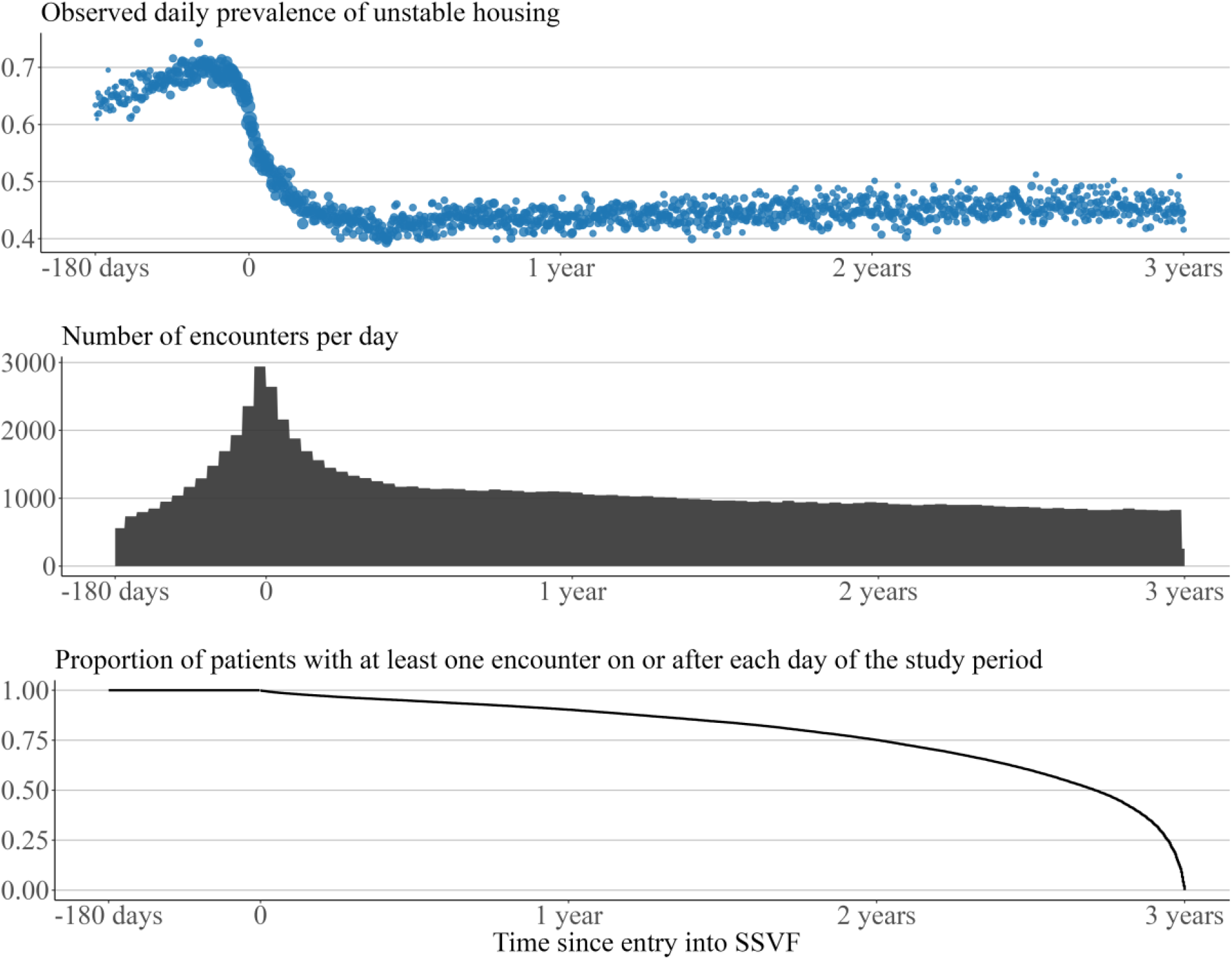
Descriptive plots of NLP classifications, number of encounters per day, and proportion of patients with at least one encounter after each day.

### Inverse Intensity Weighting

Coefficients for variables related to the encounter process from the IIW Cox proportional hazards model are shown in **Table 2**. Intensity ratios (IR) greater than 1 indicate that a variable is associated with an increased risk of having a housing-related encounter while intensity ratios less than 1 indicate a lower risk of an encounter. Being unstably housed during their previous housing-related encounter was associated with an elevated risk of visiting (IR = 1.29, 95% CI = [1.29, 1.30]), supporting our initial hypothesis that unstably housed patients are observed more frequently and that the patient’s housing history should be adjusted for. Being admitted to an inpatient service during the previous encounter was strongly associated with an increased risk (4.14 [4.12, 4.16]), as did being enrolled in SSVF on the previous day (2.49 [2.47, 2.51]). More frequent encounters (1.03 [1.03, 1.03]) and a higher proportion of unstable encounters (1.10 [1.09, 1.11]) in the six months before entering SSVF was associated with a slightly increased risk of visiting, as was a reported history of homeless in the last three years (1.08 [1.08, 1.09]), suggesting that patients with a longer history of housing instability at baseline have a higher probability of engaging in housing-related services over the following three years.

**Table 2.** Coefficient estimates of the proportional intensity model.

| **Variable** | **IR<sup>†</sup>** | **95% CI<sup>†</sup>** |
| --- | --- | --- |
| Last encounter classified as unstable | 1.29 | 1.29, 1.30 |
| Inpatient admission during last encounter | 4.14 | 4.12, 4.16 |
| Enrolled in SSVF on previous day | 2.49 | 2.47, 2.51 |
| Number of encounters in 180 days before SSVF | 1.03 | 1.03, 1.03 |
| Proportion of encounters classified as unstable | 1.10 | 1.09, 1.11 |
| Homeless in last 3 years | 1.08 | 1.08, 1.09 |
| Age |  |  |
| <40 y | REF |  |
| 40 to <50 y | 1.03 | 1.02, 1.04 |
| 50 to <60y | 1.14 | 1.13, 1.14 |
| >=60 y | 1.08 | 1.07, 1.09 |
| Sex |  |  |
| Male | REF |  |
| Female | 1.00 | 1.00, 1.01 |
| Missing | 1.10 | 1.08, 1.12 |
| Race |  |  |
| White | REF |  |
| Black/African American | 0.99 | 0.98, 0.99 |
| American Indian/Alaska Native | 1.07 | 1.05, 1.08 |
| Other/Missing | 0.88 | 0.87, 0.90 |
| Total monthly income |  |  |
| 0 | REF |  |
| >0 to 500 | 1.01 | 1.00, 1.02 |
| >500 to 1500 | 0.99 | 0.98, 0.99 |
| >1500 | 0.89 | 0.88, 0.90 |
| Education |  |  |
| Less than high school | REF |  |

| Variable | IR<sup>1</sup> | 95% CI<sup>1</sup> |
| --- | --- | --- |
| High school diploma | 0.96 | 0.95, 0.97 |
| Some college | 0.94 | 0.93, 0.96 |
| College degree | 0.92 | 0.91, 0.93 |
| Missing/Unknown | 0.98 | 0.96, 0.99 |
| Spouse or partner | 0.90 | 0.90, 0.91 |
| Number of children | 0.98 | 0.98, 0.98 |
| Rurality |  |  |
| Urban | REF |  |
| Rural | 0.96 | 0.95, 0.97 |
| Missing | 0.94 | 0.92, 0.96 |
| Medicaid | 1.09 | 1.09, 1.10 |
| Medicare | 1.06 | 1.05, 1.07 |
| Mental health diagnosis | 1.13 | 1.13, 1.14 |
| VA outpatient costs in last year | 1.00 | 1.00, 1.00 |
| VA inpatient costs in last year | 1.00 | 1.00, 1.00 |
| Rapid Rehousing/Homelessness Prevention |  |  |
| RR | REF |  |
| HP | 0.98 | 0.97, 0.98 |
| Both/Missing | 1.00 | 0.99, 1.01 |
| Temporary financial assistance | 0.98 | 0.98, 0.99 |
| Case management | 0.97 | 0.97, 0.98 |
| HUD-VASH | 1.26 | 1.26, 1.27 |
| GPD | 1.04 | 1.04, 1.05 |
| Other | 1.08 | 1.07, 1.09 |

<sup>1</sup>IR = Intensity Ratio, CI = Confidence Interval

Risk of having a housing-related encounter was also related to several demographic variables. Compared to patients without any income, patients with an income greater than $1,500/month had a decreased risk of visiting (0.89 [0.88, 0.90]). Patients with higher levels of education tended to visit less frequently compared to patients without a high school diploma (IR for college degree = 0.92 [0.91, 0.93]). Rural patients were slightly less likely to visit than urban patients (0.96 [0.95, 0.97]). Having a spouse or partner was associated with a lower risk of having an encounter (0.90 [0.90, 0.91]), as was having larger number of children (IR of 0.98 for each additional child). Veterans who were engaged in other homelessness services and were receiving additional services and follow up through programs such as HUD-VASH (1.26 [1.26, 1.27]) had a higher risk of an encounter.

### Outcome regression models

**Figure 2.** shows the predicted probability of unstable housing each day for the entire cohort both with and without IIW. Similar to the crude counts, the weighted model shows a dramatic decline in housing instability immediately following entry into SSVF, decreasing from 0.583 (95% Confidence Band = [0.571, 0.596]) on day 0 to 0.395 ([0.383, 0.408]) by day 90. After that point, housing instability began to increase slightly but remained below the original prevalence by the final day (0.399 [0.387, 0.412]). Comparing the weighted model to the unweighted model shows that IIW resulted in a similar trend but consistently lower estimates of housing instability. This difference is smallest at the beginning of the study period (absolute difference on Day 0 = 0.013), which corresponds to the time when patients were visiting most frequently as shown in **Figure 1b**. The difference increased in later parts of the study when visits were more infrequent, with an absolute difference on day 1095 of 0.058.

**Figure 2.**
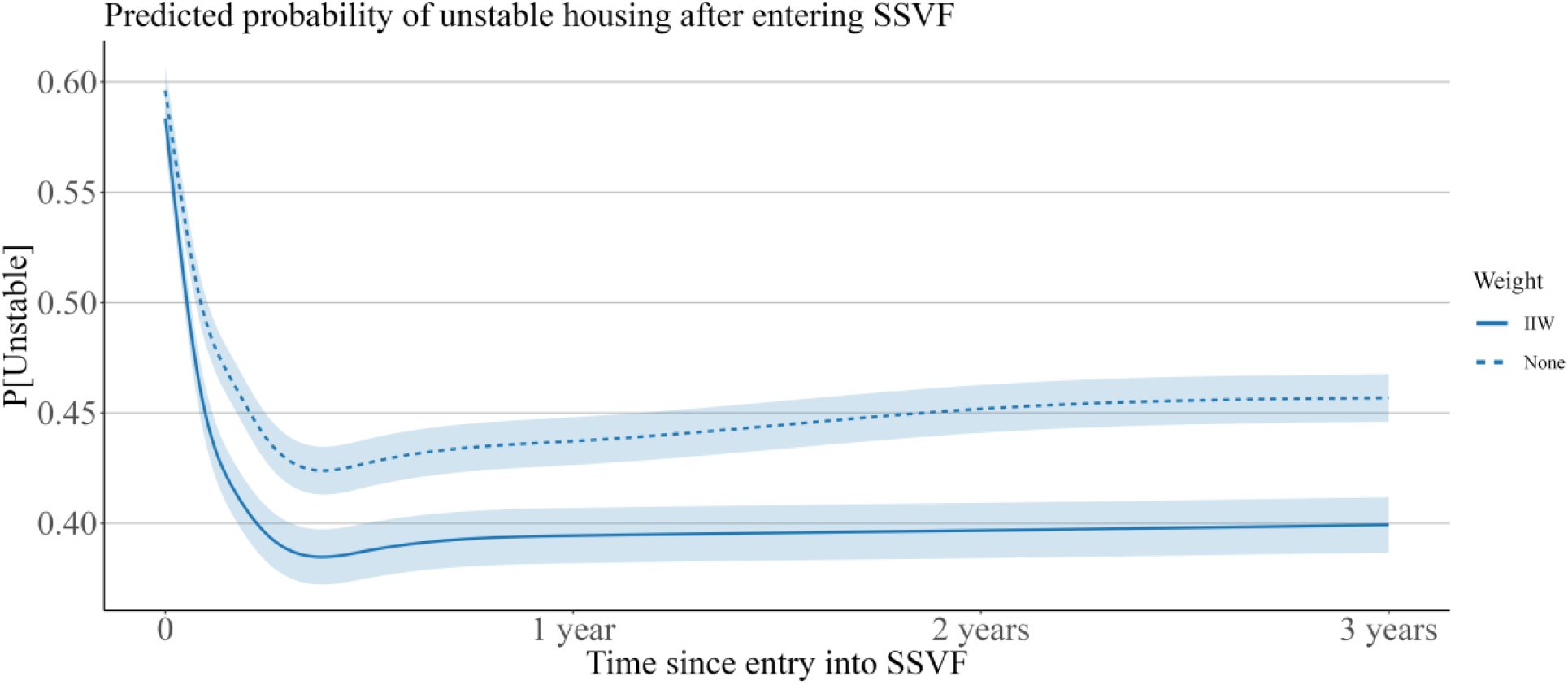
Weighted and unweighted estimated probabilities and 95% confidence band of unstable housing after entering SSVF.

## Discussion

In this work, we performed a longitudinal analysis of housing instability using EHR data. Using NLP, we collected repeated measures of patient-level housing status over long periods of time for a large cohort of homeless-experienced Veterans and compared the probability of unstable housing between patients receiving different forms of housing assistance. There was a large degree of encounter irregularity and incompleteness of data. Unstably housed patients appear to have had more frequent housing-related encounters and are thus overrepresented in the dataset. This makes intuitive sense in the context of this cohort, as patients who were unstably housed were more likely to engage with the VA and have housing information documented in the EHR. Using inverse intensity weighting in the regression model led to reduced estimates of housing instability. The difference between crude and adjusted estimates suggests that failing to account for the underlying encounter process could lead to positively biased estimates of housing instability. Inverse intensity weighting, which has been used previously in randomized trials with irregular follow up but rarely with observational EHR data, offers a promising method for reducing this bias.

This study represents an important step in methods for studying long-term housing outcomes. Traditional methods of collecting longitudinal housing outcomes studies include prospective surveys, ^20,21^ which are expensive to administer over time. Few studies have used observational EHR data to study housing instability longitudinally. A number of previous studies have used NLP to identify housing instability in clinical notes, but they did not attempt to repeatedly measure patient housing status over time.^5–10^ In VA, some studies have used the results of a standardized homelessness screener administered to Veterans nationally to examine follow up housing outcomes for Veterans with a prior positive screen.^22–25^ These studies are limited by the relatively infrequent use of the homelessness screener by clinicians, with only a small proportion of patients having at least one repeat screening.^24^ Our findings suggest that clinical notes contain more frequent documentation housing status and that leveraging this information can lead to improved measurement of housing status over time. Additionally, while the homelessness screener has been adopted nationally in the VA healthcare system, our approach may be more easily adapted to other healthcare systems and EHRs^26^ and could also be applied to other social determinants of health that are documented in clinical notes.

The results of the inverse intensity model also provide unique insights into patient characteristics associated with increased engagement with housing-related services. Several socioeconomic factors (e.g., education, income, household structure, rurality) were associated with the probability of having an encounter related to housing. These observed relationships may be due to a combination of these variables being risk factors for continued housing instability, which in turn would lead to more frequent housing-related visits over time, and underlying differences in access to care or engagement with the VA healthcare system. While this analysis was restricted to encounters with a housing-related note, this methodology could be applied in other settings to study factors associated with engaging with other services or to identify discrepancies in healthcare access.

Future work will apply the methods described here to evaluate the effectiveness of housing interventions such as SSVF. Previous work examining the effect of SSVF interventions or risk factors for SSVF enrollees have been limited to studying housing outcomes at the time patients exited SSVF.^15,16^ Additionally, these methods have broad applicability beyond the domain of housing instability and are relevant to any study examining longitudinal outcomes with EHR data. Documentation of a health condition depends on a patient interacting with the healthcare system and on subsequent documentation. This will rarely, if ever, happen in regular intervals and is highly likely to result in differential degrees of incomplete data depending on the patient’s disease status. Addressing bias due to informative and irregular collection of outcomes is thus an important step in any longitudinal analysis that uses EHR data.

This study has limitations. First, we did not account for possible measurement error due to NLP misclassification. While previous work demonstrated that ReHouSED is more accurate than structured administrative data, measurement error is present in our outcome variable. Future work should incorporate models that account for uncertainty in the outcomes due to misclassification. Second, inferences made using IIW are based on the untestable assessment at random assumption. Sensitivity analyses should be used in future work to determine the impact of violations to this assumption on analysis results. Third, we performed this analysis in the setting of the VA and using data from a cohort of Veterans who enrolled in a homelessness program. The findings here may not generalize to other subgroups of patients or to data from other healthcare systems.

## Conclusion

The EHR is a rich data source for studying long-term outcomes of patients who have experienced homelessness. NLP can be used to extract information from clinical texts and build a longitudinal dataset of repeated measures of housing instability. However, documentation in the EHR depends on patients presenting for care and is thus highly irregular and prone to incomplete information. Adjusting for encounter irregularity using inverse intensity weighting can reduce bias due to incomplete information and informative assessment times. Future work will apply these methods to evaluate interventions targeting homelessness in the VA.

## Data Availability

Due to the sensitive and protected nature of the data used in this study, data is not available to be shared.

## Footnotes

* https://www.hudexchange.info/programs/hmis/

* https://github.com/abchapman93/rehoused_longitudinal

‡ https://epullenayegum.github.io/IrregLong/

## References

1. Hatef E, Rouhizadeh M, Tia I, Lasser E, Hill-Briggs F, Marsteller J, et al. Assessing the availability of data on social and behavioral determinants in structured and unstructured electronic health records: A retrospective analysis of a multilevel health care system. JMIR Med Inform. 2019;7(3):1–15.

2. Patra BG, Sharma MM, Vekaria V, Adekkanattu P, Patterson O v., Glicksberg B, et al. Extracting social determinants of health from electronic health records using natural language processing: a systematic review. J Am Med Inform Assoc [Internet]. 2021 Dec 1 [cited 2022 Mar 6];28(12):2716. Available from: /pmc/articles/PMC8633615/

3. Gundlapalli A v., Carter ME, Palmer M, Ginter T, Redd A, Pickard S, et al. Using natural language processing on the free text of clinical documents to screen for evidence of homelessness among US veterans. AMIA. Annual Symposium proceedings / AMIA Symposium AMIA Symposium. 2013;2013:537–46.

4. Chapman AB, Jones A, Kelley AT, Jones B, Gawron L, Montgomery AE, et al. ReHouSED: A novel measurement of Veteran housing stability using natural language processing. J Biomed Inform [Internet]. 2021;122:103903. Available from: https://doi.org/10.1016/j.jbi.2021.103903

5. Lybarger K, Dobbins NJ, Long R, Singh A, Wedgeworth P, Ozuner O, et al. Leveraging Natural Language Processing to Augment Structured Social Determinants of Health Data in the Electronic Health Record. 2022;1–23. Available from: http://arxiv.org/abs/2212.07538

6. Conway M, Keyhani S, Christensen L, South BR, Vali M, Walter LC, et al. Moonstone: A novel natural language processing system for inferring social risk from clinical narratives. J Biomed Semantics. 2019 Apr 11;10(1).

7. Lybarger K, Yetisgen M. The 2022 n2c2/UW Shared Task on Extracting Social Determinants of Health. 2022;1–25.

8. Gundlapalli A V., Carter ME, Palmer M, Ginter T, Redd A, Pickard S, et al. Using natural language processing on the free text of clinical documents to screen for evidence of homelessness among US veterans. AMIA. Annual Symposium proceedings / AMIA Symposium AMIA Symposium. 2013;2013:537–46.

9. Mitra A, Pradhan R, Melamed RD, Chen K, Hoaglin DC, Tucker KL, et al. Associations Between Natural Language Processing–Enriched Social Determinants of Health and Suicide Death Among US Veterans. JAMA Netw Open [Internet]. 2023 Mar 1 [cited 2023 Mar 15];6(3):e233079–e233079. Available from: https://jamanetwork.com/journals/jamanetworkopen/fullarticle/2802468

10. Hatef E, Rouhizadeh M, Nau C, Xie F, Rouillard C, Abu-Nasser M, et al. Development and assessment of a natural language processing model to identify residential instability in electronic health records’ unstructured data: a comparison of 3 integrated healthcare delivery systems. JAMIA Open [Internet]. [cited 2022 Dec 19];5(1):1–10. Available from: https://doi.org/10.1093/jamiaopen/ooac006

11. Pullenayegum EM, Scharfstein DO. Randomized Trials with Repeatedly Measured Outcomes: Handling Irregular and Potentially Informative Assessment Times. Epidemiol Rev. 2022;44(17):121–37.

12. Lokku A, Birken CS, Maguire JL, Pullenayegum EM. Summarizing the extent of visit irregularity in longitudinal data. International Journal of Biostatistics. 2021;1–9.

13. Lin H, Scharfstein DO, Rosenheck RA. Analysis of Longitudinal Data with Irregular, Outcome-Dependent Follow-Up. Journal of the Royal Statistical Society Series B [Internet]. 2004;66(3):791–813. Available from: https://www.jstor.org/stable/3647506

14. Pullenayegum EM, Lim LSH. Longitudinal data subject to irregular observation: A review of methods with a focus on visit processes, assumptions, and study design. Stat Methods Med Res. 2016;25(6):2992–3014.

15. Nelson RE, Byrne TH, Suo Y, Cook J, Pettey W, Gundlapalli A v., et al. Association of Temporary Financial Assistance With Housing Stability Among US Veterans in the Supportive Services for Veteran Families Program. JAMA Netw Open. 2021 Feb 10;4(2):e2037047.

16. Byrne T, Montgomery AE, Chapman AB, Pettey W, Effiong A, Suo Y, et al. Predictors of homeless service utilization and stable housing status among Veterans receiving services from a nationwide homelessness prevention and rapid rehousing program. Eval Program Plann [Internet]. 2023 Apr 1 [cited 2023 Jan 1];97:102223. Available from: https://linkinghub.elsevier.com/retrieve/pii/S014971892200177X

17. Eyre H, Chapman AB, Peterson KS, Shi J, Alba PR, Jones MM, et al. Launching into clinical space with medspaCy: a new clinical text processing toolkit in Python. AMIA Annual Symposium Proceedings [Internet]. 2021 [cited 2023 Mar 16];2021:438. Available from: /pmc/articles/PMC8861690/

18. Charlson ME, Pompei P, Ales KL, MacKenzie CR. A new method of classifying prognostic comorbidity in longitudinal studies: development and validation. J Chronic Dis [Internet]. 1987 [cited 2023 Mar 15];40(5):373–83. Available from: https://pubmed.ncbi.nlm.nih.gov/3558716/

19. Bůžková P, Lumley T. Longitudinal Data Analysis for Generalized Linear Models with Follow-up Dependent on Outcome-Related Variables. Academy of Management Review. 2006;31(2):386–408.

20. Gubits D, Shinn M, Wood M, Bell S, Dastrup S, Solari CD, et al. Family Options Study 3-Year Impacts of Housing and Services Interventions for Homeless Families Family Options Study 3-Year Impacts of Housing and Services Interventions for Homeless Families 3-Year Impacts of Housing and Services Interventions for Homele. 2016;

21. Tsai J, Jones N, Szymkowiak D, Rosenheck RA. Longitudinal study of the housing and mental health outcomes of tenants appearing in eviction court. Soc Psychiatry Psychiatr Epidemiol [Internet]. 2021;56(9):1679–86. Available from: https://doi.org/10.1007/s00127-020-01953-2

22. Montgomery AE, Fargo JD, Byrne TH, Kane V, Culhane DP. Universal screening for homelessness and risk for homelessness in the Veterans Health Administration. Am J Public Health. 2013;103(SUPPL. 2):210–2.

23. Montgomery AE, Rahman FAKM, Cusack M, Varley A, Byrne T. Correlates of Transitions into Housing Instability among Veterans Accessing Veterans Health Administration Health Care. Med Care. 2020;58(12):1105–10.

24. Byrne T, Fargo JD, Montgomery AE, Roberts CB, Culhane DP, Kane V. Screening for Homelessness in the Veterans Health Administration: Monitoring Housing Stability through Repeat Screening. Public Health Reports. 1974;130(6):684–92.

25. Montgomery AE. Understanding the Dynamics of Homelessness among Veterans Receiving Outpatient Care: Lessons Learned from Universal Screening. Annals of the American Academy of Political and Social Science. 2021 Jan 1;693(1):230–43.

26. Chapman AB, Peterson KS, Rutter E, Nevers M, Zhang M, Ying J, et al. Development and evaluation of an interoperable natural language processing system for identifying pneumonia across clinical settings of care and institutions. JAMIA Open [Internet]. 2022 [cited 2023 Jan 1];5(4). Available from: https://doi.org/10.1093/jamiaopen/ooac114

